# A Pilot Report on Extracting Symptom Onset Date and Time from Clinical Notes in Patients Presenting with Chest Pain

**DOI:** 10.1101/2024.12.26.24319658

**Authors:** Anjaly George, Aashrith Maisa, Caitlin Dreisbach, Sukardi Suba

**Affiliations:** Goergen Institute for Data Science, University of Rochester; School of Nursing, University of Rochester

**Keywords:** clinical notes, electronic health records, natural language processing, temporality, time expression

## Abstract

Acute coronary syndrome (ACS) is an acute heart disease that often evolves rapidly. In ACS patients presenting with no-ST-segment elevation (NSTE-ACS), the timing of symptom onset pre-hospital may inform the disease stage and prognosis. We pilot-tested two off-the-shelf natural language processing (NLP) pipelines, namely *parsedatetime* and *regular expression* (*regex*), to extract date and time (DateTime) information of patient-reported chest pain symptoms from electronic health records (EHR) clinical notes. We included three types of clinical notes (N=71): History and Physical (n=49), Emergency Department Screening (n=3), and Triage Notes (n=19). All notes were manually annotated for the true DateTime of symptom onset. *Parsedatetime* returned matching DateTime outputs in 36 notes (50.7%), while *regex* returned zero matched outputs. *Parsedatetime* performed better than *regex*, although it was still suboptimal. Both pipelines require constant refinement and custom improvements. Methods for a large-scale, automated DateTime extraction from EHR clinical notes further investigation.

## Introduction

Natural language processing (NLP) techniques offer automated solutions for identifying and understanding unstructured text in medical research, particularly for data extraction from clinician notes in electronic health records (EHRs). While previous studies have demonstrated the utility of NLP in identifying patient symptoms from clinical notes,^1^ most implementations do not address the timing of symptom onset, a key dimension of symptom documentation. Identifying symptom onset is important in both clinical and research contexts because it provides essential context about the timing and progression of disease processes. The onset of symptoms marks the beginning of a disease’s clinical course or an acute change in clinical status and plays a critical role in diagnosis, treatment, and prognostication. Traditional methods, such as manual chart review, to determine this information are resource-intensive, time-consuming, and cost-prohibitive, particularly when scaled for quality improvement initiatives, institutional audits, or research studies.

For acute medical conditions, such as acute coronary syndrome (ACS), stroke, or sepsis, the precise time of symptom onset is crucial for timely decision-making and intervention, as delays in treatment are directly associated with worsened patient outcomes. Of particular interest, ACS often begins with patient-reported chest pain, which can be described in various word descriptors (e.g., dull, pressure, stabbing, heaviness, or squeezing).^2^ These symptoms reflect the pathophysiological process of myocardial damage, most commonly due to blockage of coronary arteries.^2-4^ The timing of chest pain or other anginal equivalent symptoms helps clinicians determine the urgency of care and eligibility for specific time-sensitive therapies such as percutaneous coronary intervention (PCI).^5-7^ The well-established “door-to-balloon time” metric underscores the importance of minimizing delays between hospital arrival and treatment,^5-7^ but pre-hospital delays—determined by the time between symptom onset and hospital arrival—are also opportunities to optimize outcomes^6,8,9^. Without precision in measuring symptom onset, the ability to evaluate and mitigate these delays is compromised.

For this present Brief Report, our primary intent was to extract the date <u>and</u> time (or *DateTime*) of symptom onset from EHR clinical notes when a patient described having symptoms before their hospital arrival. Previous methods have mostly focused on extracting *Date* information^10,11^, but, to our knowledge, reports on extracting DateTime variables have been nonexistent.

## Methods

To extract the DateTime of symptom onset from clinical notes, we tested several existing off-the-shelf NLP methods available in Python. Specifically, we evaluated three methods—*spaCy, parsedatetime*, and *regular expressions*—by adopting previously documented pipelines^10^ to two randomly selected clinical notes with as minimal modification as possible. This preliminary step aimed to optimize our limited resources (i.e., student research assistant time, effort, and computational capacity) by focusing on pipelines with the greatest likelihood of success. Each method was qualitatively assessed (SS and AM) by manually reviewing its ability to detect and extract relevant time expressions from the notes. Using an exclusion approach, we eliminated one pipeline (*spaCy*) and proceeded with the two better-performing methods, *parsedatetime* and *regular expressions* (*regex*), for further refinement and testing. During this phase, we also incorporated rules to infer symptom onset in cases where exact times were not explicitly stated in the clinical notes, following the American Heart Association (AHA) key data elements for chest pain^2^ and coronary revascularization^12^ (see **Table 1**).

**Table 1.** Rules for unspecified symptom onset time.

| Expression | Recommended Time |
| --- | --- |
| *Rules as recommended by the AHA key data elements guidelines* |  |
| “morning” | 07:00 |
| “lunch time” | 12:00 |
| “afternoon” | 15:00 |
| “dinner time” | 18:00 |
| “evening” | 22:00 |
| “awaken from sleep” | 03:00 |
| If there are indications that chest pain symptoms occurred more than once (intermittent), use the most recent ischemic symptom onset prior to hospital presentation. |  |
| *Additional rules not specified by the AHA guidelines.* |  |
| “midmorning” | 09:00 |
| “wake up in the morning” | 06:00 (*assuming the general population wake-up time is 6 AM*) |
| “after lunch” | 13:00 (*assuming it takes approximately an hour for a mealtime*) |
| “after dinner” | 19:00 (*assuming it takes approximately an hour for a mealtime*) |
| 1 or 2 or 3 or 4 days ago | Same hour time with 24-hour increment backward.<br><u>Example:</u> For a note with DateTime 01/01/2000 08:35, if it described the symptom started “1 day ago,” the onset DateTime would be 12/31/1999 08:35, and so on. |
| If the notes provide a time range, use the closest value to the notes' DateTime. | Assuming the symptom is intermittent, take the most recent time (mimicking the AHA rule above). <u>Examples:</u><br>• “chest pain or pressure began around 0200-0300” → use the second value “0300”.<br>• “2-3 days ago” → use “2 days ago”<br>• “3-5 weeks ago” → use “3 weeks ago” |

*Parsedatetime*^13^ utilizes the `parsedatetime` library to improve the precision and efficiency of temporal data extraction from clinical notes and allows parsing human-readable date and time expressions into DateTime objects. The method begins by initializing a `parsedatetime` calendar object and then applies the `nlp` function to parse and identify specific DateTime expressions within the clinical text, using a designated reference date. It also can interpret various date and time phrases, including relative dates and times; for example, “yesterday,” “next week,” “four hours ago,” or dates like “03/20.” These are commonly expressed in clinical documentation. For example, in a clinical note, “*c/o cp which began last night at 9 pm while he was watching TV, denies being diaphoretic or nausea. has taken 6 nitro since last night. has had 6 stents in past. ekg in triage*,” the function would detect “*last night at 9 pm*” as a temporal phrase. After detecting time expressions, it uses its built-in functionality to convert the time expression into a standard format based on the note date time reference.

In cases where the initial parsing fails to detect relevant expressions, a fallback method is employed to ensure comprehensive extraction. This approach iteratively parses text while focusing on explicit time mentions, such as numeric expressions like “*4:45 am*.” Regular expressions are used to identify explicit time phrases, which are then converted to a standardized DateTime format by inferring missing components (e.g., AM/PM). For instance, in a clinical note, “*66 y/o male presents to SMH ED with complains of chest pain since morning 4*.*45 am*,” the fallback method recognizes and corrects the “.” symbol in “*4*.*45 am*” to “:” and successfully captures “*4:45 am*” as the temporal expression. By combining advanced and fallback parsing mechanisms, *parsedatetime* ensures robust handling of both explicit and contextual temporal information within clinical notes.

*Regular expression (regex)*^14^ is a useful syntax that can be implemented through libraries or modules to identify and extract date and time patterns from texts based on predefined patterns and specific formats for dates and times (e.g., “Mar 2, 2016,” “1 AM,” or “21:15”). It also allows other variations and contexts in natural language, such as relative dates (e.g., “today” or “last night”), month names (e.g., “Jan 3”), or prepositions (e.g., “on” or “at”). With pattern matching, *regex* scripts can help isolate date and time strings in the clinical notes, extract them, and convert them into DateTime object format.

In this *regex* method, we developed a suite of six regular expressions to accurately parse and identify temporal expressions and varied time formats in clinical notes data. Each regular expression was tailored to capture specific structures, such as “am/pm” formats, 24-hour military time, and relative terms like “morning” or “afternoon.” Compiling each expression with case-insensitive matching ensured broad coverage of temporal information across different notations and conventions. During parsing, the method iterates through each clinical note, systematically applying these predefined expressions to extract relevant time markers, which are then stored for further analysis. For instance, in a clinical note, “*C/o midsternal chest pain radiating to right arm and neck since 0530 today*,” the phrase “0530 today” would be detected as a time expression.

### Data input and model modification

The clinical notes used in this dataset were extracted by the URMC Clinical & Translational Science Institute (CTSI) Informatics Team and provided to us in .*xlsx* format, with column variables containing patients’ information, the DateTime of each note, and the clinical notes. We selected and tested 71 clinical notes from 71 individual patients: H&P (n=49), ED Screening – First Contact (n=3), and Triage Notes (n=19). Two expert clinicians (SS and CD) manually reviewed and annotated all 71 notes for the true DateTime of the chest pain onset, following the AHA key data elements for chest pain^2^ and coronary revascularization^12^. Both annotators have a clinical background and extensive experience with chart review for research and patient care, which is necessary to perform the annotation. We did not perform any data manipulation of the texts, such as removing symbols, editing jargon or abbreviations, or relabeling notes sections or headers. We also did not pre-process and label section headers of the note texts like in other previous studies^10^. Because the H&P and ED Screening notes had a preset structure or sub-sections, we focused on selecting brief descriptions or summary sentences where clinicians would describe the patient-reported history of present illness and symptoms (see examples **Figure 1** below). We extracted these brief description sentences of the notes and then used them as inputs to the NLP methods. For the Triage Notes, we included all sentences within the notes. We refined our codes to include as many DateTime expressions as possible, informed by our experiences with conducting manual chart reviews. We evaluated the NLP methods quantitatively for the total match outputs to the true DateTime and qualitatively for the output format and mismatch. The code used for this pilot can be found on the project’s GitHub: https://github.com/suksuba/datetime-expression. All study procedures have been approved by the University of Rochester Institutional Review Board (STUDY00008105).

**Figure 1.**
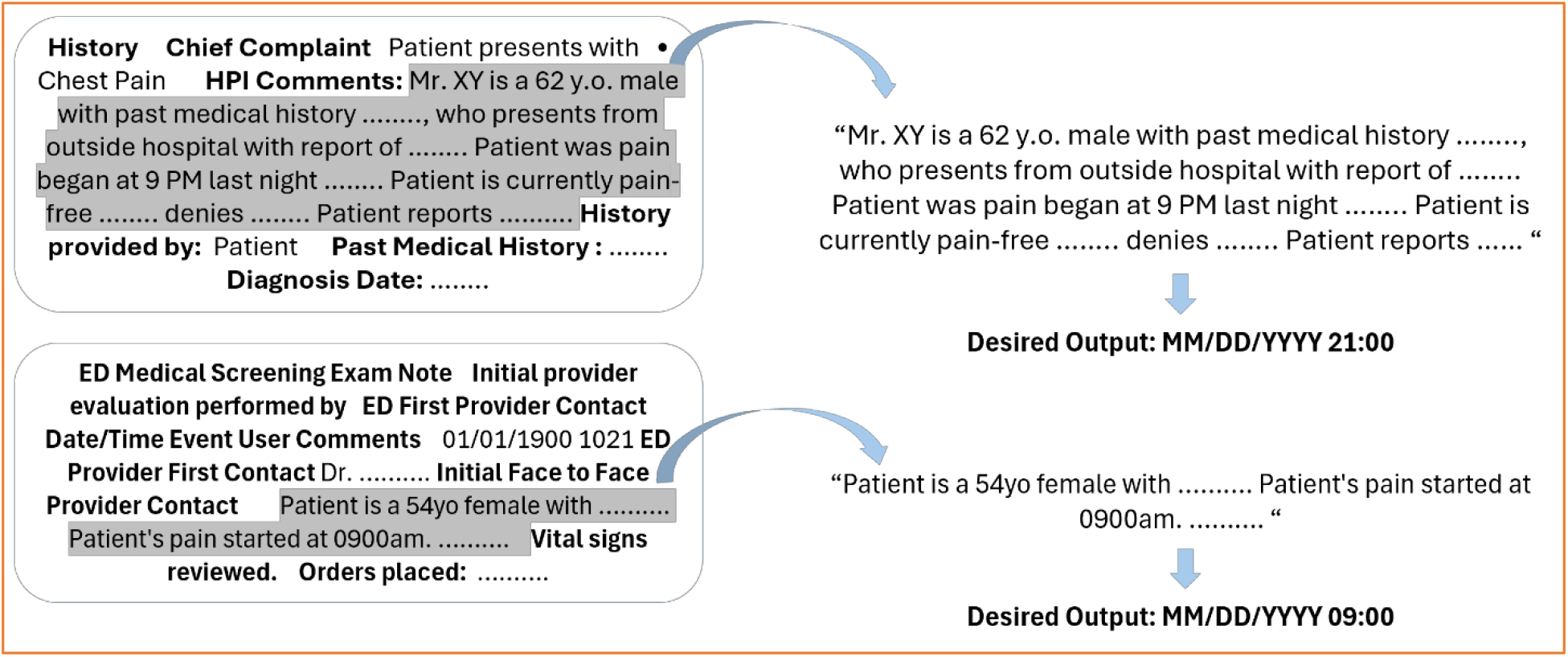
Examples of H&P (top) and ED Screening notes (bottom). Both notes have preset sections/sub-sections (bolded). We extracted descriptions of patient-reported chest pain complaints (greyed) and used the sentences as input into the NLP pipelines. The desired outputs of DateTime are shown in a standard MM/DD/YYYY HH:MM format. The date, time, and age above are de-identified.

## Results

In 71 clinical notes, the parsedatetime pipeline produced one or more DateTime outputs (up to eight) for chest pain onset, with the highest number of outputs generated for H&P notes. Upon reviewing the outputs for accuracy—defined as DateTime outputs in the ‘MM/DD/YYYY HH:MM’ format correctly matching the true symptom onset DateTime based on expert annotation—the pipeline failed to produce any matches in 35 notes (49.3%). In the remaining 36 notes (50.7%), the pipeline successfully generated one accurate output. The distribution of DateTime outputs for each category (zero matches and one match) is detailed in **Table 2**.

**Table 2.** Distribution of notes and outputs returned by the *parsedatetime* pipeline.

| Results | Total Notes<br>(N=71) | H&P<br>(n=49) | Triage<br>(n=19) | ED Screening<br>(n=3) |
| --- | --- | --- | --- | --- |
| Zero matched | 35 (49.3%) | 23 (46.9%) | 11 (57.9%) | 1 (33.3%) |
| One output | 15 | 6 | 8 | 1 |
| Two outputs | 12 | 9 | 3 | - |
| Three outputs | 2 | 2 | - | - |
| Four outputs | 3 | 3 | - | - |
| Five outputs | 1 | 1 | - | - |
| Seven outputs | 1 | 1 | - | - |
| Eight outputs | 1 | 1 | - | - |
| One matched | 36 (50.7%) | 26 (53.1%) | 8 (42.1%) | 2 (66.7%) |
| One output | 13 | 8 | 4 | 1 |
| Two outputs | 9 | 7 | 1 | 1 |
| Three outputs | 8 | 7 | 1 | - |
| Four outputs | 4 | 2 | 2 | - |
| Six outputs | 1 | 1 | - | - |
| Seven outputs | 1 | 1 | - | - |

**Note:** The number of outputs (zero to eight) indicates the total DateTime returned by the pipeline for each note. Each output was manually reviewed to determine if it matched the true date and time of chest pain onset from expert annotation.

The *regex* pipeline returned no DateTime outputs in the expected ‘MM/DD/YYYY HH:MM’ format across all notes. In seven notes (9.9%), it failed to detect any date/time expressions entirely. In the remaining 64 notes (90.1%), the pipeline identified at least one date/time expression; however, these outputs were limited to formats such as “date,” “time,” partial dates (e.g., year only), or relative expressions. The distribution of these results is summarized in (**Table 3**).

**Table 3.** Expressions format and examples returned by the *regex* pipeline.

| Extracted date/time expressions: 97 total |  |  | Examples |
| --- | --- | --- | --- |
| Format | HH:MM | 16 | 01:30, 02:45, 20:30 |
|  | HH AM/PM | 23 | 10 pm, 11pm , 3am, 4am , 11:30 am |
|  | HHMM (Military time) | 13 | 0100, 0630, 0045 |
|  | Hour Time + relative expression<br>(e.g., this morning, today, last evening) | 24 | 2200 yesterday, 11:30 pm last night, 3am today, 9 pm last night, 0315 today |
|  | YYYY | 21 | 2004 , 2008 |

## Discussion

Our symptom onset extraction from 71 clinical notes showed that *parsedatetime* has somewhat low-intermediate accuracy, while *regex* fails to provide any correct output in extracting the DateTime information we set to test. *Parsedatetime* may potentially be more useful than *regex* for automated DateTime extraction. Our findings support researchers in considering alternative methods for identifying the DateTime of symptom onset.

*Parsedatetime*—despite being effective for standard temporal expressions—occasionally misidentifies unrelated numerical values, such as medication doses or historical counts (e.g., “6 mg” or “6 stents”), as time expressions. This misclassification poses potential challenges for downstream analysis, likely affecting the pipeline into failing to recognize temporal time expressions. Our pilot test showed that this pipeline required more extensive computational tasks than anticipated (e.g., extensive code development, pipeline requiring broader rules or linguistic expressions of time), especially considering we tested it only on 71 notes. The observed errors (zero matched and multiple DateTime outputs) underscore a limitation of *parsedatetime* when parsing the complex and highly variable language in clinical notes, suggesting a need for refined filtering or supplementary methods to exclude non-temporal numerical data accurately. One particularly noteworthy finding we noted was the pipeline’s inability to recognize the contextual relationship between a time expression and its elaboration in a sentence where the symptom onset time was explicitly stated: “*He was in his usual state of health until approx 3AM today, when he says he woke and had severe chest discomfort/pressure*.” While the pipeline correctly identified “*3AM today*” and “*he woke up*” (see rules in **Table 1**) as time expressions in this Triage Note, it failed to understand that the expression “*he woke up*” was an elaboration of the “*3AM*” time. Consequently, the pipeline returned two identical DateTime outputs for this note.

Similarly, *regex* seems like it should be effective for the task. However, it relies on a relatively large number of regular expressions to capture the broad linguistic variability of clinical language, which may present limitations for larger datasets or when language patterns shift unexpectedly. Modifying the method, we used various time expressions commonly used in the EHR clinical notes. However, we realized it would require us to review a larger number of existing clinical notes manually to be able to scope most of the temporal expressions used by clinicians, which seems counterintuitive to our goal to avoid manual tasks by utilizing automated methods. The variability and complexity of temporal language require ongoing refinement of regular expressions, which may reduce scalability and adaptability. As illustrated in the analysis above, the method’s reliance on predefined scenarios can complicate accurate temporal extraction when faced with unexpected or unstructured expressions, a potential drawback for processing extensive clinical datasets.

To our knowledge, this is the first study that tests the application of NLP methods to extract symptom onset DateTime from clinical notes. Our pilot test using *parsedatetime* and *regex* methods failed to extract DateTime at the accuracy level we hoped it would achieve. Our pilot test was built on the previous work by Fu et al.^10^, in which they tested the same NLP methods in extracting *Dates* from clinical notes in patients with myeloproliferative neoplasms. However, their task was to extract Date information only, the format of which is generally simpler than the DateTime expression. Dates are generally expressed explicitly in clinical notes, in full or partial, such as *Month DD, YYYY, MM/DD/YYYY, MM/DD/YY, MM/DD, MM/YY*, or *Month DD*. They are also often expressed as a narrative relative, such as *today, last month*, or *6 years ago*. We found similar expressions in our data but sought to extract the additional specific *HH:MM time* to the dates. Due to its manageable number, we manually reviewed and tabulated the NLP methods for accuracy, allowing us to judge its performance qualitatively. As Fu et al. demonstrated,^10^ we also found that both NLP methods are suboptimal in executing our task for automated DateTime extraction.

As an alternative to NLP methods, large language models (LLMs) may offer significant advantages for extracting symptom onset date and time from clinical notes due to their adaptability and ability to handle variability in language. Clinical notes often describe symptom onset using unstructured, variable, or implicit language (e.g., “*yesterday morning*,” “*while at work two days ago*”), which LLMs can interpret by leveraging their extensive pretraining on diverse datasets. Unlike traditional methods, like the ones used in this study, that rely on manual feature engineering and require large, annotated datasets, LLMs excel in few-shot (i.e., performing a new task after being exposed to only a small number of examples) and zero-shot (i.e., ability to perform on a task it hasn’t learned explicitly) learning, making them well-suited for clinical domains where labeled data are scarce. Their ability to perform temporal reasoning, infer implicit information, and handle noisy or inconsistent text further enhances their utility. Additionally, LLMs are scalable and adaptable, allowing integration of multiple tasks, such as extracting symptom onset, duration, and severity, with minimal reconfiguration. Our team is currently testing the ability of existing LLMs to extract symptom onset DateTime.

We noted some limitations in our pilot testing, all due to the limited resources we faced. We had a small number of samples (N=71 notes) in this initial pilot test. However, this number of annotated clinical notes is relatively similar to that in other studies on NLP technique application,^10,15,16^ and it falls within the sample size range commonly reported in pilot or feasibility studies^17^. We did not perform pre-processing of notes structure or section headers because our focus was to pilot-test the performance of the NLP methods. Instead, we manually cleaned up the notes to make them “analysis-ready,” which was doable due to the small number of clinical notes we annotated and tested.

### Application and usability for nursing research and quality improvement

The application of automated DateTime extraction systems extends into nursing research and quality improvement, offering significant utility for advancing patient outcome research and evidence-based care. In nursing research, such systems provide a scalable and efficient method for collecting temporal data, enabling large-scale studies on the relationship between symptom onset and patient outcomes, disease progression, care timelines, or treatment efficacy. This facilitates the identification of trends and care gaps, contributing to developing interventions that improve patient safety and outcomes. For quality improvement initiatives, automated extraction tools support the evaluation of institutional performance metrics, such as time-to-intervention or adherence to care protocols, by providing consistent and accurate data from electronic health records. These capabilities enhance the rigor of nursing research and empower nurses to drive system-level changes that elevate the quality and efficiency of care delivery.

## Conclusion

We pilot-tested two off-the-shelf NLP pipelines, *parsedatetime* and *regex*, for an automated DateTime extraction task. *Parsedatetime* performed better than *regex* but was, alas, still suboptimal. Both pipelines’ computational costs—encompassing human resources, code development, and time—can escalate significantly due to the need for constant refinement and custom improvements. These challenges are particularly pronounced in research institutions, where limited resources often necessitate balancing innovation with efficiency. Scaling these pipelines for large-scale applications remains difficult, underscoring the need for further investigation into developing an optimal automated method for DateTime extraction.

## Data Availability

Codes and outputs produced in the present work are contained in the manuscript. Clinical notes cannot be publicly shared as they are represent potentially identifying and sensitive patient data.

https://github.com/suksuba/datetime-expression

## Corresponding author

*Sukardi Suba, PhD, RN* 601 Elmwood Ave. Box SON, Rochester, NY 14642

## Acknowledgment

We thank the CTSI Clinical Informatics team for their assistance in curating the electronic health record data for this study.

## CRediT

**Anjaly George** and **Aashrith Maisa** contributed to the data analysis, methodology, validation, writing – review and editing. **Anjaly George** also contributed to writing – the original draft. **Caitlin Dreisbach** contributed expert review and writing for the final manuscript. **Sukardi Suba** contributed to the conceptualization, data curation, data analysis, funding acquisition, methodology, project administration, resources, supervision, data validation, and writing and reviewing the original draft and editing revisions.

## References

1. Koleck TA, Dreisbach C, Bourne PE, Bakken S. Natural language processing of symptoms documented in free-text narratives of electronic health records: a systematic review. J Am Med Inform Assoc. Apr 1 2019;26(4):364–379. doi:10.1093/jamia/ocy173

2. Anderson HVS, Masri SC, Abdallah MS, et al. 2022 ACC/AHA Key Data Elements and Definitions for Chest Pain and Acute Myocardial Infarction: A Report of the American Heart Association/American College of Cardiology Joint Committee on Clinical Data Standards. Circ Cardiovasc Qual Outcomes. Oct 2022;15(10):e000112. doi:10.1161/HCQ.0000000000000112

3. Thygesen K, Alpert JS, Jaffe AS, et al. Fourth Universal Definition of Myocardial Infarction (2018). J Am Coll Cardiol. Oct 30 2018;72(18):2231–2264. doi:10.1016/j.jacc.2018.08.1038

4. Zegre-Hemsey JK, Burke LA, DeVon HA. Patient-reported symptoms improve prediction of acute coronary syndrome in the emergency department. Res Nurs Health. Oct 2018;41(5):459–468. doi:10.1002/nur.21902

5. Byrne RA, Rossello X, Coughlan JJ, et al. 2023 ESC Guidelines for the management of acute coronary syndromes. Eur Heart J. Oct 12 2023;44(38):3720–3826. doi:10.1093/eurheartj/ehad191

6. Lawton JS, Tamis-Holland JE, Bangalore S, et al. 2021 ACC/AHA/SCAI Guideline for Coronary Artery Revascularization: A Report of the American College of Cardiology/American Heart Association Joint Committee on Clinical Practice Guidelines. Circulation. Jan 18 2022;145(3):e18–e114. doi:10.1161/CIR.0000000000001038

7. Kontos MC, de Lemos JA, Deitelzweig SB, et al. 2022 ACC Expert Consensus Decision Pathway on the Evaluation and Disposition of Acute Chest Pain in the Emergency Department: A Report of the American College of Cardiology Solution Set Oversight Committee. J Am Coll Cardiol. Nov 15 2022;80(20):1925–1960. doi:10.1016/j.jacc.2022.08.750

8. De Luca G, Suryapranata H, Ottervanger JP, Antman EM. Time delay to treatment and mortality in primary angioplasty for acute myocardial infarction - Every minute of delay counts. Circulation. Mar 16 2004;109(10):1223–1225. doi:10.1161/01.Cir.0000121424.76486.20

9. De Luca G, Suryapranata H, Zijlstra F, et al. Symptom-onset-to-balloon time and mortality in patients with acute myocardial infarction treated by primary angioplasty. Journal of the American College of Cardiology. Sep 17 2003;42(6):991–997. doi:10.1016/S0735-1097(03)00919-7

10. Fu JT, Sholle E, Krichevsky S, Scandura J, Campion TR. Extracting and classifying diagnosis dates from clinical notes: A case study. J Biomed Inform. Oct 2020;110:103569. doi:10.1016/j.jbi.2020.103569

11. Wang L, Wampfler J, Dispenzieri A, Xu H, Yang P, Liu H. Achievability to Extract Specific Date Information for Cancer Research. AMIA Annu Symp Proc. 2019;2019:893–902.

12. Dehmer GJ, Badhwar V, Bermudez EA, et al. 2020 AHA/ACC Key Data Elements and Definitions for Coronary Revascularization: A Report of the American College of Cardiology/American Heart Association Task Force on Clinical Data Standards (Writing Committee to Develop Clinical Data Standards for Coronary Revascularization). Circ Cardiovasc Qual Outcomes. Apr 2020;13(4):e000059. doi:10.1161/HCQ.0000000000000059

13. Tylor M. Parse human-readable date/time strings. 2024. https://github.com/bear/parsedatetime

14. Bird S. timex.py. 2024. https://github.com/nltk/nltk_contrib/blob/master/nltk_contrib/timex.py

15. Lindvall C, Deng CY, Moseley E, et al. Natural Language Processing to Identify Advance Care Planning Documentation in a Multisite Pragmatic Clinical Trial. J Pain Symptom Manage. Jan 2022;63(1):e29–e36. doi:10.1016/j.jpainsymman.2021.06.025

16. Petch J, Batt J, Murray J, Mamdani M. Extracting Clinical Features From Dictated Ambulatory Consult Notes Using a Commercially Available Natural Language Processing Tool: Pilot, Retrospective, Cross-Sectional Validation Study. JMIR Med Inform. Nov 1 2019;7(4):e12575. doi:10.2196/12575

17. Totton N, Lin J, Julious S, Chowdhury M, Brand A. A review of sample sizes for UK pilot and feasibility studies on the ISRCTN registry from 2013 to 2020. Pilot Feasibility Stud. Nov 21 2023;9(1):188. doi:10.1186/s40814-023-01416-w

